# Acceptance of approaching death in cancer and non-cancer patients according to general practitioners; a European Study in Primary Care

**DOI:** 10.64898/2026.04.23.26351589

**Authors:** Myrte Zijlstra, Natasja Raijmakers, Nienke Veldhuijzen, Lieve Van den Block, Sarah Moreels, Tomas Vega Alonso, Guido Miccinesi, Bregje Onwuteaka-Philipsen

## Abstract

**Background:** Acceptance of death is an important aspect of a ‘good death’ and an indicator of high-quality palliative care. Limited evidence exists on the extent to which patients accept their approaching death and which socio-demographic or end-of-life care characteristics are associated with acceptance, in both cancer and non-cancer patients.

**Methods:** We conducted a retrospective cross-sectional survey in representative GP networks in the Netherlands and Italy (2013–2015), and Belgium and Spain (2013–2014). GPs registered all deceased adult patients in their practice, reporting health and care characteristics in the last three months of life and the level of acceptance of approaching death on a four-point scale (“1. Yes, completely” to “4. No, not at all”). Non-sudden deaths were included, totaling 2,796 patients (1,474 cancer; 1,322 non-cancer).

**Results:** Acceptance was recorded for 97% of patients (n=2,713), of which 17% were assessed as ‘unknown’. GPs assessed that 30% of patients had complete acceptance, with similar proportions in cancer and non-cancer patients (30% vs 29%). Multivariable logistic regression showed that older age (OR 1.03, 95%CI 1.02–1.04, p<0.01), country (OR 0.27 Belgium, OR 0.11 Italy, OR 0.10 Spain; reference: Netherlands), and palliative care by the GP until death (OR 1.39, 95%CI 1.07–1.79, p<0.01) were independently associated with complete acceptance.

**Conclusion:** Socio-demographic, contextual, and end-of-life care factors influence GP-assessed acceptance of approaching death, whereas a cancer diagnosis does not. These findings emphasize the importance of culturally sensitive, age-appropriate, and palliative care–oriented approaches to support patient acceptance at the end of life.

## Introduction

Regardless of how or where we are born, the fact that everyone eventually dies unites people across all cultures. The importance of high-quality end-of-life care is being increasingly recognized. In light of adequate end-of-life care, quality of dying and death is currently recognized as an important objective. Research on Quality of Death (QoD) is increasing, with particular emphasis on the measurability of the concept and its inherent subjectivity.^[1, 2]^ Quality of Death comprises and identifies several aspects and determinants in the literature, including socio-demographic and cultural patient characteristics and end-of-life care elements such as place of death, aggressive end-of-life-care and end-of-life communication.^[3-5]^

To assess QoD from the perspective of the patients and relatives, Downey et al examined the underlying domains of the Quality of Dying and Death (QODD) questionnaire, which include symptom control, connectedness, preparation and transcendence.^[6]^ Acceptance of death is associated with both preparation for and transcendence of mortality.^[7]^ It is defined as the rational acknowledgment of one’s inevitable death, combined with the positive emotional integration of this realization.^[8]^ Acceptance of approaching death may function as a coping mechanism for terminally ill patients, providing self-reassurance as they come to terms with their limited time and difficult circumstances.^[9]^ This process may also be conceptualized as the counterpart of death anxiety. Both death acceptance and death anxiety are associated with several outcome measures. Previous studies have shown that patients who were cognitively aware of their terminal status and who felt “at peace” showed more positive outcomes, such as lower rates of anticipatory grief and psychological distress, and higher rates of advance care planning than patients who were not at peace.^[10-12]^ Moreover, acceptance of death is associated with higher perceived quality of care by family caregivers.^[13]^ As such, acceptance of death is seen as an important aspect of a “good death” and considered an important outcome indicator of high-quality palliative care.

Within Europe, attitudes toward life and death vary between countries.^[14]^ Known factors associated to acceptance of death include older age, religious beliefs, cultural influences, psychological wellbeing (anxiety and depression, coping), health status, advanced disease, and preparedness.^[8, 11, 15-18]^ For patients with cancer, the predictability of the disease course may lead to a greater sense of preparedness for death.^[19]^ In addition, the more widespread integration of palliative care in oncology may further contribute to this increased preparedness and acceptance of death, differentiating patients dying of cancer from those dying of non-cancerous diseases.^[20]^ On the other hand, cancer patients often report high levels of perceived injustice, which may hinder acceptance and increase psychological distress.^[21]^

General practitioners are well positioned to assess patients’ acceptance of an approaching death due to their central role in palliative care, longstanding relationships with patients, and involvement in end-of-life decision-making.^[22]^ This study aimed to assess the level of acceptance of an approaching death in cancer and non-cancer patients in Europe, as perceived by their general practitioners, and to explore which socio-demographic and end-of-life care characteristics are associated with this acceptance.

## Methods

### Setting and design

This study was based on data collected as part of the European Sentinel General Practitioner Networks Monitoring End-of-Life Care (EURO-SENTIMELC) study, an ongoing cross-national, retrospective mortality follow-back study with representative general practitioner (GP) networks in Belgium, the Netherlands, Italy (Tuscany region) and Spain (Castile and León and Valencian Community regions).^[23]^ GPs from these countries collected information on demographic, health and several end-of-life care characteristics of each deceased adult patient in the practices of these GP networks, using a standardized questionnaire. In Belgium, the Netherlands and Spain, existing GP sentinel networks for epidemiological surveillance participated in the study, while in Italy, a network was created specifically for the study. To avoid selecting GPs with a special interest in end-of-life care, recruited GPs were not informed about the content of the surveillance before participation. The data used in this study were collected from 01/01/2013 to 31/12/2014 in Belgium and Spain, from 01/01/2013 to 31/12/2015 in the Netherlands, and from 01/06/2013 to 31/12/2015 in Italy, and accessed on 03/08/2018.

### Study population

A total of 5,449 deaths were recorded by the GP networks during the study period. As this study focuses on the degree of acceptance of an approaching death from the perspective of the GP, people whose longest place of stay in the last year of life was within the GP’s remit were selected for this analysis: people who lived at home or in a care home. Patients whose death was sudden and those with severe dementia were excluded to allow assessment of the level of acceptance of an approaching death, resulting in a total number of 2,796 patients for this analysis.

### Informed Consent, Patient Anonymity, and Ethical Approval

All participating GPs provided written informed consent at the start of each registration year after being fully informed about the study objectives and procedures. Strict protocols safeguarded patient anonymity: each patient received an anonymous reference code, and identifying patient and GP details were replaced by aggregated or coded variables.

The study was approved by the Ethical Review Board of Brussels University Hospital (Vrije Universiteit Brussel) and the Local Ethical Committee “Comitato Etico della Azienda U.S.L. n. 9 di Grosseto”, Tuscany. In the Netherlands and Spain, no additional ethical approval was required due to the retrospective and fully anonymized nature of the data.

### Measures

Within one week of reporting a patient’s death, participating GPs were asked to fill in a questionnaire.

#### Acceptance of an approaching death

The extent of the patient’s acceptance of an approaching death was assessed by the GP using a four-point scale (“1. yes, completely”, “2. yes, for the most part”, “3. no, not entirely”, “4. no, not at all”). GPs could also indicate “unknown” if they felt unable to assess the extent of acceptance.

#### Patient’s and end-of-life care characteristics

Using a standardized registration form consisting of structured and closed-ended items, GPs recorded patients’ demographic, health and end-of-life care characteristics: age at death, gender, longest place of residence in the last year of life, palliative care provided by GP, multidisciplinary palliative care initiative involved in the last three months of life, dementia, main diagnosis/cause of death, and whether or not death was sudden and unexpected.

### Statistical analyses

Descriptive statistics were used to analyze patient characteristics, end-of-life care and level of acceptance of approaching death. Logistic regression analysis was performed to assess the association between patient and care characteristics and complete acceptance of approaching death, first using univariate analysis. For this purpose, the original four-point acceptance scale was dichotomized: a score of 1 was defined as complete acceptance, and scores 2–4 as non-complete acceptance. Cases with unknown acceptance were excluded to allow for a clear comparison between complete and non-complete acceptance. The following characteristics were included in the logistic regression analysis: age, gender, country, cancer diagnosis, longest place of residence in the last year of life, palliative care provided by the GP, and involvement of a multidisciplinary (specialist) palliative care initiative. All characteristics significantly associated with complete acceptance in the univariate analyses were subsequently included in a multivariable logistic regression model, with two-sided p-values of <0.1. Due to differences in palliative care initiatives in the participating countries, an interaction effect was added (country * multidisciplinary palliative care initiative). A threshold of p < 0.05 was used for the final results of the multivariable analysis. A separate analysis was conducted to examine the association between patient and care characteristics and whether the level of acceptance of an approaching death could be assessed. Both univariate and multivariable logistic regression analyses were performed for this purpose. All statistical analyses were performed using STATA (release 17.0, StataCorp, College Station, Texas, USA).

## Results

### Characteristics of study population

A total of 2,796 deceased patients were included in the analysis, of whom 53% (n=1,474) had cancer as their primary diagnosis or cause of death (Table 1). The most common non-cancer cause of death was cardiovascular disease (17% of the total sample; 37% of the non-cancer group). The median age of the total population was 82 years (range 20-107), 49% was male, 42% lived in Belgium, and 82% lived at home or with family for most of the past year. To provide further context, study population characteristics were also examined by country (Tables A–D in S1 Table). Differences were observed in end-of-life care characteristics across countries. GP palliative care was more prevalent in the Netherlands compared to other countries (B 58% N 83% I 65% E 54%), while multidisciplinary (specialist) palliative care was less common in both the Netherlands and Italy (B 62% N 28% I 40% E 85%).

**Table 1:** Characteristics of study population, subdivided by cancer or no cancer diagnosis.

|  | All patients<br>N=2796 | Cancer patients<br>N=1474 | Non-cancer<br>patients<br>N=1322 |
| --- | --- | --- | --- |
| **<u>Sociodemographic characteristics</u>** |  |  |  |
| Age, median (range) | 82 (20-107) | 76 (27-103) | 86 (20-107) |
| Age group, n (%) |  |  |  |
| <65 yrs | 373 (13%) | 307 (21%) | 66 (5.0%) |
| 65-84 yrs | 1316 (47%) | 856 (58%) | 460 (35%) |
| ≥85 yrs | 1098 (39%) | 306 (21%) | 792 (60%) |
| Gender, n (%) |  |  |  |
| Male | 1355 (49%) | 800 (54%) | 555 (42%) |
| Female | 1428 (51%) | 671 (46%) | 757 (58%) |
| Country, n (%) |  |  |  |
| Belgium | 1163 (42%) | 595 (40%) | 568 (43%) |
| The Netherlands | 812 (29%) | 508 (34%) | 304 (23%) |
| Italy | 515 (18%) | 208 (14%) | 307 (23%) |
| Spain | 306 (11%) | 163 (11%) | 143 (11%) |
| **<u>End-of-life care characteristics</u>** |  |  |  |
| Longest place of residence last year of life, n (%) |  |  |  |
| At home/with family | 2289 (82%) | 1348 (91%) | 941 (71%) |
| Care home | 507 (18%) | 126 (8.6%) | 381 (29%) |
| Palliative care by GP, n (%) |  |  |  |
| Yes, until death | 1399 (52%) | 812 (57%) | 587 (45%) |
| Yes, but not until death | 389 (14%) | 257 (18%) | 132 (10%) |
| No | 925 (34%) | 348 (25%) | 577 (45%) |
| Multidisciplinary (specialist) palliative care initiative involved last 3 months of life, n (%)* |  |  |  |
| Yes | 1286 (51%) | 844 (62%) | 442 (38%) |
| No | 1252 (49%) | 516 (38%) | 736 (62%) |

Missings ≤5% are not reported
\* Missings 8.7% (cancer 7.2% vs non-cancer 10%)

### Acceptance of an approaching death

The level of acceptance of an approaching death was filled in for 97% (n=2,713) of the patients (n=1,417 cancer, n=1,296 non-cancer). In the remaining 3%, this assessment was missing for unspecified reasons. In the 2,713 patients for whom assessment was completed, the GP recorded the level of acceptance of an approaching death as complete in 30% (n=802) of all deceased patients—30% of cancer patients and 29% of non-cancer patients (Figure 1). The degree of complete acceptance varied across countries, with 24% in Belgium, 52% in the Netherlands, 13% in Italy, and 19% in Spain.

In 17% of the cases, acceptance was assessed by the GP as “unknown”, with country-level proportions ranging from 5% to 27% (B 21% N 5% I 21% E 27%), and this was more frequent among non-cancer patients (24%) compared to cancer patients (12%). Unknown acceptance was associated with living outside the Netherlands, not having cancer, having (mild) dementia, and not receiving palliative care from the GP (all p<0.001; Table in S2 Table). Cases with unknown acceptance were excluded from further analyses (see Methods).

**Figure. 1.**
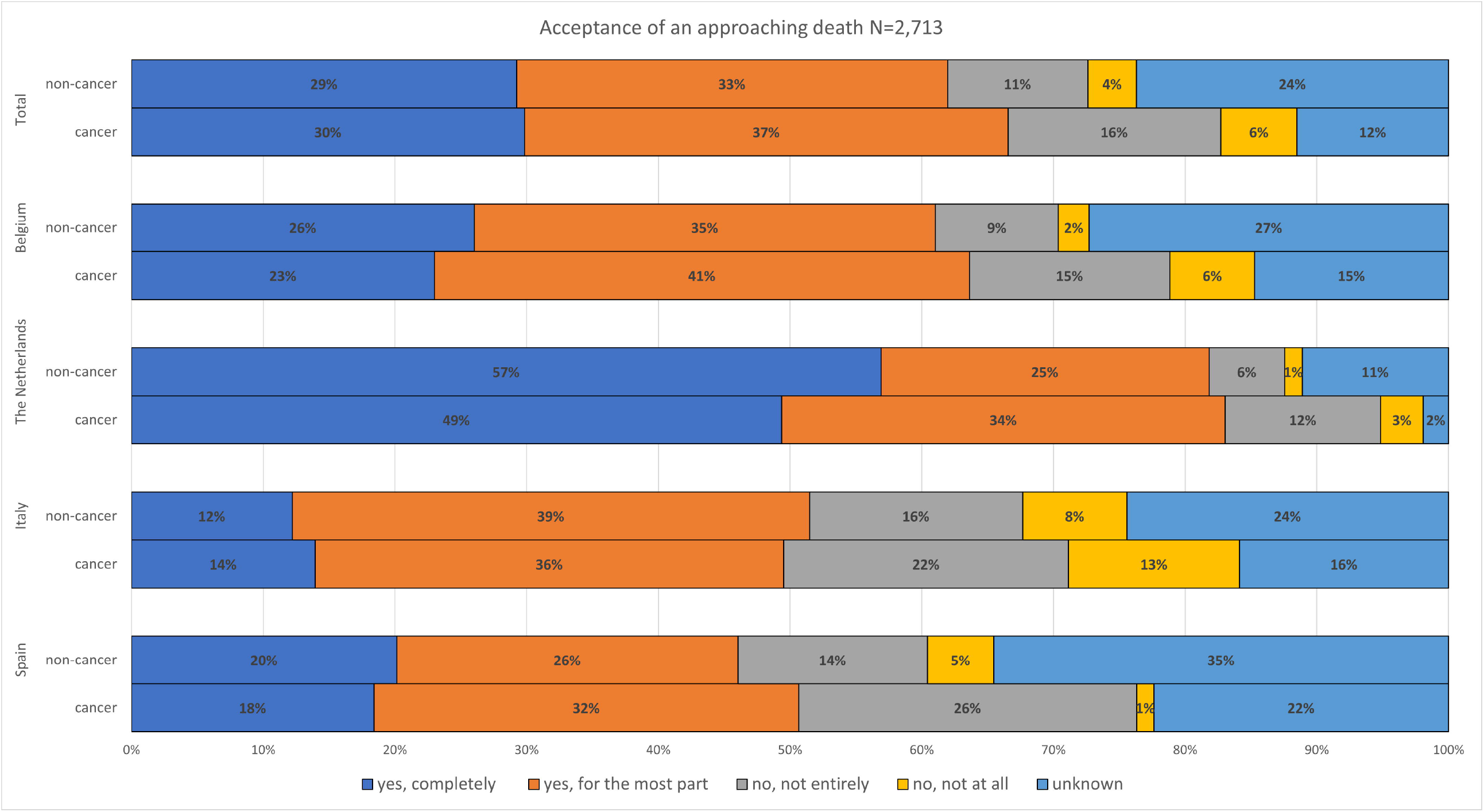
Acceptance of an approaching death by country

### Factors associated with acceptance of an approaching death

Univariate analysis showed that the following factors were associated with complete acceptance of an approaching death: (increasing) age, female gender, country, cancer diagnosis, residence in a care home, palliative care provided by a GP, and no involvement of a multidisciplinary (specialist) palliative care initiative, (all p<0.05; Table 2).

**Table 2:**
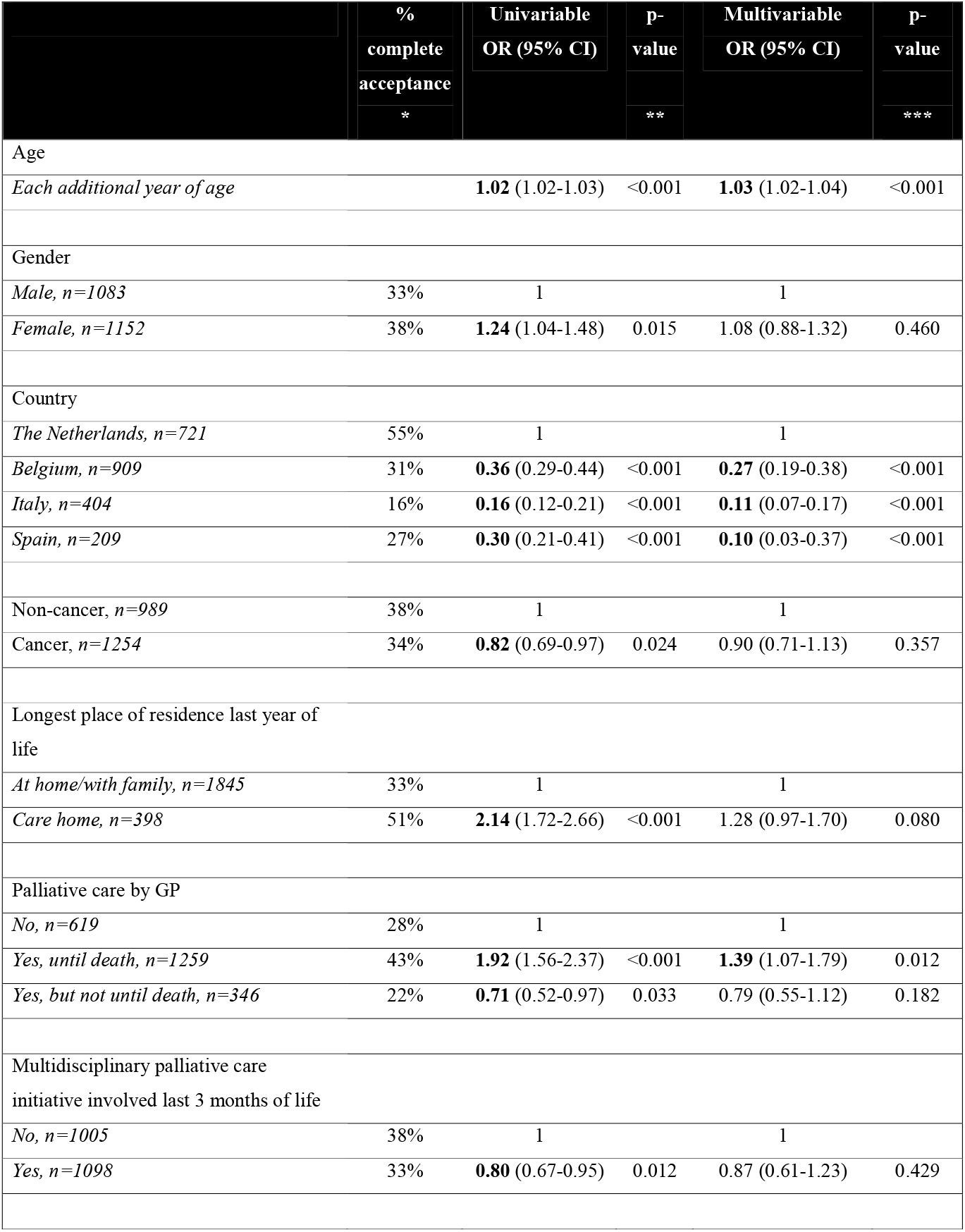
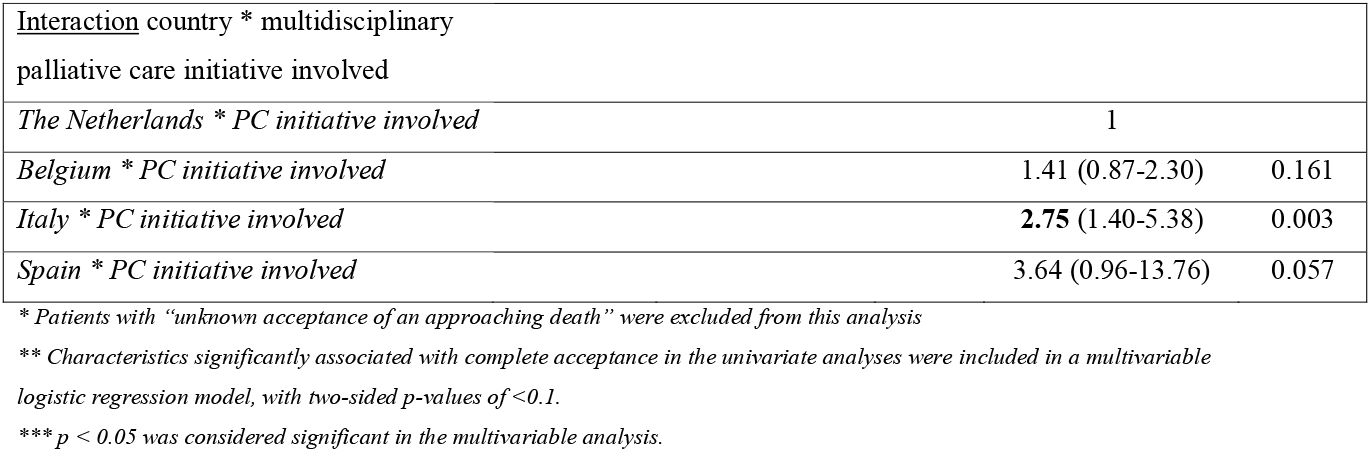
Factors associated with **complete** acceptance of approaching death.

Multivariable logistic regression analysis showed that older age was associated with having complete acceptance with their approaching death (OR 1.03 95%CI 1.02-1.04, p<0.01) (Table 2). Patients who received palliative care from their GP until death were also more likely to have complete acceptance (OR 1.39 95%CI 1.07-1.79, p<0.01). Patients were less likely to be assessed as having complete acceptance with an approaching death when living in Belgium (OR 0.27 95%CI 0.19-0.38, p<0.01), Italy (OR 0.11 95%CI 0.07-0.17, p<0.01) or Spain (OR 0.10 95%CI 0.03-0.37, p<0.01), compared to patients living in the Netherlands. In Italy, the involvement of a multidisciplinary (specialist) palliative care initiative was associated with having complete acceptance of an approaching death (OR 2.75 95%CI 1.40-5.38, p<0.01, reference country the Netherlands). Similar results were found in Spain, although not significant (OR 3.64 95%CI 0.96-13.76, p<0.057, reference country The Netherlands).

## Discussion

This European study provides valuable insights into the level of acceptance of an approaching death by patients, as assessed by general practitioners, and the factors involved in this process in four European countries. For 30% of all deceased patients, GPs indicated “complete acceptance of an approaching death” both for patients with and without cancer, but with significant differences between countries. In addition, patients of older age and who received palliative care provided by the GP until death were more likely to have “complete acceptance” of their death. In 17% of cases, the GP was not able to determine the patient’s level of acceptance of approaching death (classified as unknown), with notable variation between countries.

Comparison with previous studies on acceptance of death is challenging due to the diversity of measurement methods used to assess acceptance of death. In addition, existing studies are based on patient self-reports, whereas the present study reflects general practitioners’ perspectives. The percentage of patients in our study who reported complete acceptance of approaching death was notably lower than the 74% of patients with advanced cancer who accepted their terminal prognosis, as reported by Thompson et al.^[24]^ However, it is important to note that accepting a terminal prognosis and accepting an approaching death are not the same. The acceptance of a terminal prognosis may involve a more gradual process of adjustment over time, whereas accepting approaching death may be more immediate and confronting. In a previous study on death acceptance involving cancer patients using the Life Attitude Profile - Revised (LAP-R) acceptance subscale, with responses ranging from ‘strongly disagree’ to ‘strongly agree,’ most participants reported being ‘undecided’ (4.33/7) about the level of death acceptance they experienced, reflecting a more moderate level of acceptance, with the mean score being slightly lower than that of the normative population and patients with other chronic illnesses.^[11]^ In another study on death acceptance involving cancer patients and healthy controls, higher age, but not having cancer, was associated with higher levels of death acceptance, which is consistent with the results of our study.^[15]^ In our univariate analyses, cancer patients appeared to have lower acceptance of death; however, this difference disappeared in multivariable analyses when accounting for care characteristics and patient factors, indicating that the apparent effect of cancer diagnosis on death acceptance is mediated by these variables. In our study, we found no difference in death acceptance between men and women. Some previous studies have indicated that women experience higher levels of death anxiety, but also report no gender differences in death acceptance.^[25, 26]^ Tong et al. demonstrated that individuals with varying levels of death anxiety use different coping strategies, with those experiencing lower anxiety either accepting or avoiding death.^[27]^ These findings suggest that the relationship between death anxiety and death acceptance is more nuanced than a simple dichotomy.

End-of-life care practices and preferences vary across countries, reflecting differences in cultural values, healthcare systems, ethical norms, and national policies.^[28-30]^ Palliative care is strongly supported by law and policy in Belgium, the Netherlands and Spain, whereas in Italy it was only recognized as a legal right starting in 2010.^[30]^ The organization of end-of-life care also differs between these countries.^[30]^ In the Netherlands, GPs generally carry primary responsibility for end-of-life care, either independently or in collaboration with mobile palliative care teams. In Belgium and Spain, care is more often shared with specialized palliative home care teams. In Italy, multidisciplinary home teams predominantly provide palliative care, with GPs coordinating within community-based teams but not hospital settings.

As outlined above, GP involvement in end-of-life care varies across countries. Consistent with previous research, this study shows that GPs in the Netherlands are more strongly involved in palliative care than in the other countries, and specialist palliative care services are used less often.^[31]^ Additionally, compared to clinical specialists, GPs are more likely to address psychosocial aspects and end-of-life care issues in their communications.^[32]^ Our study demonstrated an association between GP involvement until death and higher levels of acceptance of death. While this finding suggests that continuous GP involvement may play an important role in supporting patients’ acceptance, it may also reflect how the GP’s involvement shapes their perception or judgment of the patient’s acceptance of death, as GPs were the reporting source..

Communication patterns also differ across countries. In the Netherlands, end-of-life topics are discussed most frequently, followed by Belgium, with fewer discussions in Spain and Italy.^[30]^ End-of-life practices in Southern Europe are traditionally shaped by Catholic values, with a strong emphasis on hope, family involvement, and protective communication.^[33]^ Paternalism remains strong, and partial disclosure or even deception regarding terminal diagnoses still occurs.^[34]^ Such limited transparency may inhibit open dialogue about a patient’s prognosis, potentially reducing acceptance of an approaching death.

Beyond care, individual’s perspective on life and death is influenced by their worldview, often shaped by cultural values and religious beliefs.^[17, 35]^ In both Italy and Spain, Catholicism, deeply embedded in the traditions of these countries, may influence death attitudes and the way death-related topics are approached, potentially leading to observed differences in the acceptance of death compared to the other countries. While religious individuals often report higher levels of death acceptance, beliefs about the afterlife may both alleviate death anxiety and heighten fear of judgment.^[36]^ Interestingly, some studies suggest that non-religious individuals with strong convictions may also have higher death acceptance.^[25]^ As medicalization of life and death and modernization progresses, traditional religiosity has become less central, while values emphasizing individual choice and self-expression have gained prominence.^[37]^ This shift toward individualism is particularly evident in countries like the Netherlands, where personal autonomy, self-determination, and end-of-life decision-making are promoted, which may foster a greater acceptance of death.^[38]^

### Strengths & limitations

This study’s multi-country design is a key strength, providing valuable insights into end-of-life care across diverse healthcare systems and cultural contexts. The retrospective, cross-sectional design facilitated the identification of patients nearing the end of life, a task that would be difficult in a prospective study. The evaluation of death acceptance through the judgment of GPs offers a practical and efficient assessment method. However, the approach lacks the standardization and depth of tools like the LAP-R, and the outcomes are based on the physician’s judgment, which could be influenced by their personal attitudes towards death.^[39]^ In countries where end-of-life communication is less open, GPs’ ability or tendency to assess patients’ acceptance of death may also be limited. Additionally, the study did not account for factors such as religion and symptom burden, which may have provided further insight into the determinants of death acceptance. Another limitation of this study is that the data were collected over a decade ago. Changes in healthcare delivery, policy, or public attitudes since then may have influenced current practices and outcomes. To our knowledge, however, no more recent studies have examined death acceptance in a comparable context. Moreover, the topic remains highly relevant, particularly in light of ageing populations and ongoing societal and clinical discussions about end-of-life care and dying well.^[40]^ As such, our findings continue to contribute important insights into patient characteristics and contextual factors associated with death acceptance.

### Conclusion

This study provides insights into the complex dynamics of death acceptance and the important role GPs play in this process, guiding patients through the challenges of facing death. It highlights the importance of understanding cultural and systemic factors influencing death attitudes and acceptance to improve the quality of care and communication between GPs and their patients during end-of-life stages and better meet the needs of different populations. Optimizing palliative care integration remains key, as higher death acceptance is linked to GPs providing continuous care and palliative teams’ involvement, the latter especially in southern countries.

## Acknowledgments

The authors would like to thank all general practitioners and local network coordinators participating in the EURO SENTIMELC study for their invaluable contributions to data collection and support throughout the study.

## Data availability statement

Data may be obtained from a third party and are not publicly available. Data may be made available on request to the relevant national institute.

### Key points

- Complete acceptance of approaching death was reported by GP in 30% of patients, similar for cancer and non-cancer patients.
- Acceptance of approaching death rates varied significantly between countries.
- Older patients and those receiving GP-provided palliative care until death were more likely to show complete acceptance of approaching death.
- Findings highlight the importance of culturally sensitive, age-appropriate, and palliative care– oriented approaches in public health policy and practice.

## References

1. Patrick DL, Engelberg RA, Curtis JR. Evaluating the quality of dying and death. J Pain Symptom Manage. 2001;22(3):717–26.

2. Hales S, Zimmermann C, Rodin G. Review: the quality of dying and death: a systematic review of measures. Palliat Med. 2010;24(2):127–44.

3. Naya K, Sakuramoto H, Aikawa G, Ouchi A, Yoshihara S, Ota Y, et al. Family Members’ Feedback on the “Quality of Death” of Adult Patients Who Died in Intensive Care Units and the Factors Affecting the Death Quality: A Systematic Review and Meta-Analysis. Cureus. 2024;16(4):e58344.

4. Nakazawa Y, Miyashita M, Morita T, Okumura Y, Kizawa Y, Kawagoe S, et al. Dying Patients’ Quality of Care for Five Common Causes of Death: A Nationwide Mortality Follow-Back Survey. J Palliat Med. 2024.

5. Lee JJ, Long AC, Curtis JR, Engelberg RA. The Influence of Race/Ethnicity and Education on Family Ratings of the Quality of Dying in the ICU. J Pain Symptom Manage. 2016;51(1):9–16.

6. Downey L, Curtis JR, Lafferty WE, Herting JR, Engelberg RA. The Quality of Dying and Death Questionnaire (QODD): empirical domains and theoretical perspectives. J Pain Symptom Manage. 2010;39(1):9–22.

7. Wong P. Meaning Management Theory and Death Acceptance. In: Tomer, Eliason, Wong, editors. Existential and Spiritual Issues in Death Attitudes: Erlbaum; 2008.

8. Neimeyer RA, Wittkowski J, Moser RP. Psychological research on death attitudes: an overview and evaluation. Death Stud. 2004;28(4):309–40.

9. Kyota A, Kanda K. How to come to terms with facing death: a qualitative study examining the experiences of patients with terminal Cancer. BMC Palliat Care. 2019;18(1):33.

10. Ray A, Block SD, Friedlander RJ, Zhang B, Maciejewski PK, Prigerson HG. Peaceful awareness in patients with advanced cancer. J Palliat Med. 2006;9(6):1359–68.

11. Philipp R, Mehnert A, Lo C, Muller V, Reck M, Vehling S. Characterizing death acceptance among patients with cancer. Psychooncology. 2019;28(4):854–62.

12. Davis EL, Deane FP, Lyons GCB, Barclay GD. Is Higher Acceptance Associated With Less Anticipatory Grief Among Patients in Palliative Care? J Pain Symptom Manage. 2017;54(1):120–5.

13. Lee JH, Lee YJ, Park SJ, Park YM, Lee CW, Hwang SW, et al. Patient Acceptance of Death and Symptom Control/Quality of Care Among Terminal Cancer Patients Under Inpatient Hospice Care: A Multicenter Cross-Sectional Study. Am J Hosp Palliat Care. 2025:10499091251318738.

14. Bartolome-Peral E, Coromina L. Attitudes towards Life and Death in Europe: A Comparative Analysis. Sociologicky casopis / Czech Sociological Review. 2021;56(6):835–62.

15. Pinquart M, Fröhlich C, Silbereisen RK, Wedding U. Death Acceptance in Cancer Patients. OMEGA - Journal of Death and Dying. 2006;52(3):217–35.

16. Dezutter J, Soenens B, Luyckx K, Bruyneel S, Vansteenkiste M, Duriez B, et al. The role of religion in death attitudes: distinguishing between religious belief and style of processing religious contents. Death Stud. 2009;33(1):73–92.

17. Gire JT. How Death Imitates Life: Cultural Influences on Conceptions of Death and Dying. Online Readings in Psychology and Culture. 2014;6:3.

18. Tang ST, Chou WC, Chang WC, Chen JS, Hsieh CH, Wen FH, et al. Courses of Change in Good Emotional Preparedness for Death and Accurate Prognostic Awareness and Their Associations With Psychological Distress and Quality of Life in Terminally Ill Cancer Patients’ Last Year of Life. J Pain Symptom Manage. 2019;58(4):623–31 e1.

19. Murray SA, Kendall M, Boyd K, Sheikh A. Illness trajectories and palliative care. BMJ. 2005;330(7498):1007–11.

20. Kim S, Lee K, Kim S. Knowledge, attitude, confidence, and educational needs of palliative care in nurses caring for non-cancer patients: a cross-sectional, descriptive study. BMC Palliat Care. 2020;19(1):105.

21. Secinti E, Wu W, Krueger EF, Hirsh AT, Torke AM, Hanna NH, et al. Relations of perceived injustice to psycho-spiritual outcomes in advanced lung and prostate cancer: Examining the role of acceptance and meaning making. Psychooncology. 2022;31(12):2177–84.

22. Ramanayake RP, Dilanka GV, Premasiri LW. Palliative care; role of family physicians. J Family Med Prim Care. 2016;5(2):234–7.

23. Van den Block L, Onwuteaka-Philipsen B, Meeussen K, Donker G, Giusti F, Miccinesi G, et al. Nationwide continuous monitoring of end-of-life care via representative networks of general practitioners in Europe. BMC Fam Pract. 2013;14:73.

24. Thompson GN, Chochinov HM, Wilson KG, McPherson CJ, Chary S, O’Shea FM, et al. Prognostic acceptance and the well-being of patients receiving palliative care for cancer. J Clin Oncol. 2009;27(34):5757–62.

25. Sawyer JS, Brewster ME, Ertl MM. Death anxiety and death acceptance in atheists and other nonbelievers. Death Stud. 2021;45(6):459–68.

26. Soleimani MA, Bahrami N, Allen K-A, Alimoradi Z. Death anxiety in patients with cancer: A systematic review and meta-analysis. European Journal of Oncology Nursing. 2020;48:101803.

27. Tong E, Deckert A, Gani N, Nissim R, Rydall A, Hales S, et al. The meaning of self-reported death anxiety in advanced cancer. Palliat Med. 2016;30(8):772–9.

28. Gysels M, Evans N, Menaca A, Andrew E, Toscani F, Finetti S, et al. Culture and end of life care: a scoping exercise in seven European countries. PLoS One. 2012;7(4):e34188.

29. The Economist Intelligence Unit. The Quality of Death Index 2015 – Ranking palliative care across the world. London: The Economist Intelligence Unit 2015 October 2015.

30. Evans N, Costantini M, Pasman HR, Van den Block L, Donker GA, Miccinesi G, et al. End-of-life communication: a retrospective survey of representative general practitioner networks in four countries. J Pain Symptom Manage. 2014;47(3):604–19 e3.

31. Ko W, Deliens L, Miccinesi G, Giusti F, Moreels S, Donker GA, et al. Care provided and care setting transitions in the last three months of life of cancer patients: a nationwide monitoring study in four European countries. BMC Cancer. 2014;14:960.

32. Michiels E, Deschepper R, Bilsen J, Mortier F, Deliens L. Information disclosure to terminally ill patients and their relatives: self-reported practice of Belgian clinical specialists and general practitioners. Palliative Medicine. 2009;23(4):345–53.

33. Meñaca A, Evans N, Andrew EV, Toscani F, Finetti S, Gómez-Batiste X, et al. End-of-life care across Southern Europe: a critical review of cultural similarities and differences between Italy, Spain and Portugal. Crit Rev Oncol Hematol. 2012;82(3):387–401.

34. Toscani F, Farsides C. Deception, Catholicism, and Hope: Understanding Problems in the Communication of Unfavorable Prognoses in Traditionally-Catholic Countries. The American Journal of Bioethics. 2006;6(1):W6–W18.

35. Hassan BH, Fernández-Alcántara M, García-Caro MP, Ibrahim N, Eweida RS. Cross-Cultural Comparison of Older Adults’ Emotional Responses Toward Death: A Pilot Study. Res Gerontol Nurs. 2024;17(3):112–20.

36. Jong J, Robert R, Tristan P, Si-Hua C, Naomi S, and Halberstadt J. The religious correlates of death anxiety: a systematic review and meta-analysis. Religion, Brain & Behavior. 2018;8(1):4–20.

37. Rudnev M, Savelkaeva A. Public support for the right to euthanasia: Impact of traditional religiosity and autonomy values across 37 nations. International Journal of Comparative Sociology. 2018;59:002071521878758.

38. Hofstede G. Culture’s Consequences: Comparing Values, Behaviors, Institutions, and Organizations Across Nations. 2nd ed. Thousand Oaks, CA: Sage Publications; 2001.

39. Miccinesi G, Fischer S, Paci E, Onwuteaka-Philipsen BD, Cartwright C, van der Heide A, et al. Physicians’ attitudes towards end-of-life decisions: a comparison between seven countries. Soc Sci Med. 2005;60(9):1961–74.

40. Amsterdam UMC. It’s time to accept death and give it a real place in society 2022 [Available from: https://www.amsterdamumc.org/en/research/institutes/amsterdam-public-health/news/its-time-to-accept-death-and-give-it-a-real-place-in-society-1.htm.

