## Supplement 1 Table A-D & Supplement 2 Table for "Acceptance of approaching death in cancer and non-cancer patients according to general practitioners; a European Study in Primary Care"

**Supplementary information**

S1 Table A: Characteristics of study population, subdivided by cancer or no cancer diagnosis, **Belgium**

|  | All patients  N=1163 | Cancer patients  N=595 | Non-cancer patients  N=568 |
| --- | --- | --- | --- |
| Sociodemographic characteristics |  |  |  |
| Age, median (range) | 82 (20-100) | 76 (35-101) | 86 (20-106) |
| Age group, n (%) |  |  |  |
| *<65 yrs* | 152 (13%) | 118 (20%) | 34 (6.0%) |
| *65-84 yrs* | 548 (47%) | 331 (56%) | 217 (38%) |
| *≥85 yrs* | 460 (40%) | 144 (24%) | 316 (56%) |
| Gender, n (%) |  |  |  |
| *Male* | 558 (48%) | 323 (54%) | 235 (42%) |
| *Female* | 602 (52%) | 272 (46%) | 330 (58%) |
| End-of-life care characteristics |  |  |  |
| Longest place of residence last year of life, n (%) |  |  |  |
| *At home/with family* | 877 (75%) | 526 (88%) | 351 (62%) |
| *Care home* | 286 (25%) | 69 (12%) | 217 (38%) |
| Palliative care by GP, n (%) |  |  |  |
| *Yes, until death* | 544 (47%) | 310 (52%) | 234 (42%) |
| *Yes, but not until death* | 127 (11%) | 95 (16%) | 32 (5.7%) |
| *No* | 481 (42%) | 187 (32%) | 294 (53%) |
| Multidisciplinary (specialist) palliative care initiative involved last 3 months of life, n (%) |  |  |  |
| *Yes* | 687 (62%) | 427 (73%) | 260 (49%) |
| *No* | 425 (38%) | 156 (27%) | 269 (51%) |

*Missings ≤5% are not reported*

S1 Table B: Characteristics of study population, subdivided by cancer or no cancer diagnosis, **The Netherlands**

|  | All patients  N=812 | Cancer patients  N=508 | Non-cancer patients  N=304 |
| --- | --- | --- | --- |
| Sociodemographic characteristics |  |  |  |
| Age, median (range) | 78 (27-102) | 74 (27-97) | 87 (45-102) |
| Age group, n (%) |  |  |  |
| *<65 yrs* | 138 (17%) | 118 (23%) | 20 (6.6%) |
| *65-84 yrs* | 413 (51%) | 311 (61%) | 102 (34%) |
| *≥85 yrs* | 261 (32%) | 79 (16%) | 182 (60%) |
| Gender, n (%) |  |  |  |
| *Male* | 401 (50%) | 268 (53%) | 133 (42%) |
| *Female* | 408 (50%) | 239 (47%) | 169 (58%) |
| End-of-life care characteristics |  |  |  |
| Longest place of residence last year of life, n (%) |  |  |  |
| *At home/with family* | 666 (82%) | 466 (92%) | 200 (66%) |
| *Care home* | 146 (18%) | 42 (8.3%) | 104 (34%) |
| Palliative care by GP, n (%)* |  |  |  |
| *Yes, until death* | 556 (74%) | 371 (80%) | 195 (65%) |
| *Yes, but not until death* | 63 (8.3%) | 47 (10%) | 16 (5.4%) |
| *No* | 131 (17%) | 44 (9.5%) | 87 (29%) |
| Multidisciplinary (specialist) palliative care initiative involved last 3 months of life, n (%)** |  |  |  |
| *Yes* | 199 (28%) | 141 (32%) | 58 (22%) |
| *No* | 505 (72%) | 297 (68%) | 208 (78%) |

*Missings ≤5% are not reported*

** Missings 6.4% (cancer 9.1% vs non-cancer 2.0%)*

*** Missings 13 % (cancer 14% vs non-cancer 13%)*

S1 Table C: Characteristics of study population, subdivided by cancer or no cancer diagnosis, **Italy**

|  | All patients  N=515 | Cancer patients  N=208 | Non-cancer patients  N=307 |
| --- | --- | --- | --- |
| Sociodemographic characteristics |  |  |  |
| Age, median (range) | 84 (33-103) | 78 (33-99) | 87 (49-103) |
| Age group, n (%) |  |  |  |
| *<65 yrs* | 46 (9.0%) | 38 (19%) | 8 (2.6%) |
| *65-84 yrs* | 216 (42%) | 117 (57%) | 99 (33%) |
| *≥85 yrs* | 247 (49%) | 50 (24%) | 197 (65%) |
| Gender, n (%) |  |  |  |
| *Male* | 227 (45%) | 103 (50%) | 124 (41%) |
| *Female* | 281 (55%) | 103 (50%) | 178 (59%) |
| End-of-life care characteristics |  |  |  |
| Longest place of residence last year of life, n (%) |  |  |  |
| *At home/with family* | 477 (93%) | 199 (96%) | 278 (91%) |
| *Care home* | 38 (7.4%) | 9 (4.3%) | 29 (9.5%) |
| Palliative care by GP, n (%) |  |  |  |
| *Yes, until death* | 186 (37%) | 67 (32%) | 119 (40%) |
| *Yes, but not until death* | 142 (28%) | 76 (37%) | 66 (22%) |
| *No* | 177 (35%) | 64 (31%) | 113 (38%) |
| Multidisciplinary (specialist) palliative care initiative involved last 3 months of life, n (%)* |  |  |  |
| *Yes* | 189 (40%) | 139 (71%) | 50 (18%) |
| *No* | 286 (60%) | 56 (29%) | 230 (82%) |

*Missings ≤5% are not reported*

** Missings 7.8% (cancer 6.3% vs non-cancer 8.8%)*

S1 Table D: Characteristics of study population, subdivided by cancer or no cancer diagnosis, **Spain**

|  | All patients  N=306 | Cancer patients  N=163 | Non-cancer patients  N=143 |
| --- | --- | --- | --- |
| Sociodemographic characteristics |  |  |  |
| Age, median (range) | 83 (37-107) | 78 (37-103) | 89 (51-107) |
| Age group, n (%) |  |  |  |
| *<65 yrs* | 37 (12%) | 33 (20%) | 4 (2.8%) |
| *65-84 yrs* | 139 (45%) | 97 (60%) | 42 (29%) |
| *≥85 yrs* | 130 (42%) | 33 (20%) | 97 (68%) |
| Gender, n (%) |  |  |  |
| *Male* | 169 (55%) | 106 (65%) | 63 (44%) |
| *Female* | 137 (45%) | 57 (35%) | 80 (56%) |
| End-of-life care characteristics |  |  |  |
| Longest place of residence last year of life, n (%) |  |  |  |
| *At home/with family* | 269 (88%) | 157 (96%) | 112 (78%) |
| *Care home* | 37 (12%) | 6 (3.7 %) | 31 (22%) |
| Palliative care by GP, n (%) |  |  |  |
| *Yes, until death* | 103 (35%) | 64 (41%) | 39 (28%) |
| *Yes, but not until death* | 57 (19%) | 39 (25%) | 18 (13%) |
| *No* | 136 (56%) | 53 (34%) | 83 (59%) |
| Multidisciplinary (specialist) palliative care initiative involved last 3 months of life, n (%)* |  |  |  |
| *Yes* | 211 (85%) | 137 (95%) | 74 (72%) |
| *No* | 36 (15%) | 7 (4.9%) | 29 (28%) |

*Missings ≤5% are not reported*

** Missings 19% (cancer 12% vs non-cancer 28%)*

S2 Table: Factors associated with **unknown** level of acceptance of approaching death

|  | % unknown acceptance | Univariable  OR (95% CI) | p-value* | Multivariable  OR (95% CI) | p-value** |
| --- | --- | --- | --- | --- | --- |
| Age |  |  |  |  |  |
| *Each additional year of age* |  | **1.01** (1.01-1.02) | 0.001 | 1.00 (0.99-1.01) | 0.563 |
| Gender |  |  |  |  |  |
| *Male, n=1315* | 18% | 1 |  |  |  |
| *Female, n=1386* | 17% | 0.95 (0.78-1.16) | 0.602 |  |  |
| Country |  |  |  |  |  |
| *The Netherlands, n=763* | 6% | 1 |  | 1 |  |
| *Belgium, n=1148* | 21% | **4.51** (3.21-6.36) | <0.001 | **4.55** (2.96-6.99) | <0.001 |
| *Italy, n=511* | 21% | **4.55** (3.12-6.63) | <0.001 | **4.14** (2.60-6.59) | <0.001 |
| *Spain, n=291* | 28% | **6.74** (4.50-10.07) | <0.001 | **5.82** (3.42-9.91) | <0.001 |
| Non-cancer, *n=1296* | 24% | 1 |  | 1 |  |
| Cancer, *n=1417* | 12% | **0.42** (0.34-0.52) | <0.001 | **0.64** (0.49-0.84) | 0.001 |
| No dementia, *n=2229* | 14% | 1 |  | 1 |  |
| Mild dementia, *n=449* | 31% | **2.63** (2.08-3.32) | <0.001 | **2.44** (1.83-3.26) | <0.001 |
| Longest place of residence last year of life |  |  |  |  |  |
| *At home/with family, n=2220* | 17% | 1 |  |  |  |
| *Care home, n=493* | 19% | 1.17 (0.91-1.51) | 0.207 |  |  |
| Palliative care by GP |  |  |  |  |  |
| *No, n=907* | 32% | 1 |  | 1 |  |
| *Yes, until death, n=1394* | 10% | **0.23** (0.18-0.29) | <0.001 | **0.33** (0.25-0.43) | <0.001 |
| *Yes, but not until death, n=387* | 11% | **0.25** (0.18-0.36) | <0.001 | **0.31** (0.20-0.46) | <0.001 |
| Multidisciplinary palliative care initiative involved last 3 months of life |  |  |  |  |  |
| *No, n=1238* | 19% | 1 |  | 1 |  |
| *Yes, n=1278* | 14% | **0.71** (0.57-0.87) | 0.001 | 0.81 (0.62-1.06) | 0.118 |

** characteristics significantly associated with complete acceptance in the univariate analyses were included in a multivariable logistic regression model, with two-sided p-values of <0.1.*

*** p < 0.05 was considered significant in the multivariable analysis.*
